# THE PATHOGENESIS OF THROMBOEMBOLIC DISEASE IN COVID-19 PATIENTS: COULD BE A CATASTROPHIC ANTIPHOSPHOLIPID SYNDROME?

**DOI:** 10.1101/2020.04.30.20086397

**Authors:** G Previtali, M Seghezzi, V Moioli, A Sonzogni, L Cerutti, R Marozzi, R Ravasio, A. Gianatti, G. Guerra, MG Alessio

**Affiliations:** Clinical Chemistry Laboratory, ASST Papa Giovanni XXIII Hospital, Bergamo, Italy; Pathological Anatomy, ASST Papa Giovanni XXIII Hospital, Bergamo, Italy

## Abstract

**Background:** The most severely COVID-19 patients need intensive care and show increased risk of thromboembolic events. Although some patients meet the diagnostic criteria for the Disseminated Intravascular Coagulation, the pathogenesis of the diffuse thrombotic status remains unclear.

The aim of the present study is to evaluate the presence of antiphospholipid antibodies (aPL) in sera of deceased patients with autoptic proven thrombotic microangiopathy to evaluate if some patients may have developed Catastrophic Antiphospholipid Syndrome (CAPS).

**Methods:** Thirty-five patients were enrolled. The available medical history, comorbidities, therapies, laboratory and autopsy findings were collected post-mortem from clinical records.

IgA, IgG and IgM anti cardiolipin (ACA) and anti β2 glycoprotein 1 (β2GP1) antibodies, IgG and IgM anti phosphatidylserine/prothrombin (PS/PT) antibodies were tested for all the patients.

**Results:** 3/35 (8.6%) patients were slightly positive for aPL: one for ACA IgG and two for ACA IgM but values were low (< 3X the cut off). No patients tested positive for ACA IgA neither for β2GP1 isotypes. 3/35 (8.6%) patients were positive for PS/PT, one for IgG and two for IgM, but values were less than 2X the cut off. No patients showed simultaneous positivity for ACA and PS/ PT.

**Conclusions:** It is difficult to categorize the vascular events into a conventional disease: we did not find significant association with anti-phospholipid antibodies. It is most likely that several factors contribute to trigger the hypercoagulability status and the thromboembolism but, on the basis our results, CAPS is probably not involved into the pathogenesis of these phenomena.

## Introduction

In December 2019 an epidemic of pneumonia caused by SARS-CoV-2, a novel coronavirus, developed from Wuhan, Hubei Province in China [1]. The outbreak rapidly spread all over the world and the World Health Organization (WHO) declared the SARS-CoV-2 disease (named COVID-19) a pandemic on 11th March 2020. COVID-19 outbreak had quite hard impact on the Italian health system; many infected subjects are symptomless or present a flu-like disease, but about 15-20% of patients can develop a severe syndrome, characterized by interstitial pneumonia with alveolar damage and severe Acute Respiratory Distress Syndrome (ARDS), leading in 5% of cases to death [2]. The most severely patients affected by COVID-19 need intensive care and show an increased risk of thromboembolic events, since the SARS-CoV-2 infection cause an hyperinflammatory response targeting endothelium and leading to a pro-coagulation state, worsened by hypoxia and prolonged immobilization. These patients can develop pulmonary thrombosis and/or multi-organ failure due to thrombotic microangiopathy (TMA) [3–6].

Although some patients meet the diagnostic criteria for the Disseminated Intravascular Coagulation (DIC) [7, 8], there is a group of COVID 19 severely ill patients in which the pathogenesis of the diffuse thrombotic status remains unclear.

The Catastrophic Antiphospholipid Syndrome (CAPS) is a rare, life-threatening condition characterized by multiple thrombosis, affecting mainly small vessels and involving three or more organs, developing in less than a week and associated with persistent antiphospholipid antibodies (aPL) positivity. (9) Peculiarly CAPS is triggered by precipitating factors and infections, mainly in respiratory tract, are the most frequent events (10). The underlying pathogenetic mechanism is not completely understood, but could be precipitated by a “cytokine storm”, with diffuse vasculitis and direct or indirect damage on endothelium (11)

The clinical course and the autopsy findings in patients died for COVID-19 in Bergamo (the epicenter city of COVID-19 outbreak in Italy) suggest that some patients may have developed CAPS.

The aim of the present study is to evaluate the presence of aPL in sera of deceased patients with autoptic proven TMA.

## Material and Methods

### Patients

Seventy-five autopsies on patients with molecular proven infection of SARS-CoV-2 were performed at Papa Giovanni XXIII Hospital in Bergamo between 19^th^ March and 09^th^ April 2020. The autopsies were performed in Airborne Infection Isolation autopsy rooms and the personnel used the correct Personal Protection Equipment (PPE), according to “Engineering control and PPE recommendations for autopsies”. Only skilled pathologists were enrolled to perform post-mortem procedures.

7 patients infected during the hospital stay for other diseases were excluded (5 organ transplant and 2 neoplastic disease). Sixty-eight patients were eligible for this study: they were admitted to the hospital for COVID-19 and died during the hospital stay. Sera for aPL testing, collected 24h before death and frozen at −20°C, were available only for thirty-five patients that were enrolled for the study (26 male and 9 female, ratio 2.88:1; age range 57y-92y, mean and median age 73y). All the clinical records were evaluated postmortem to collect the available medical history, comorbidities, therapies, laboratory and autopsy findings.

### Laboratory testing

IgA, IgG and IgM anti cardiolipin antibodies (ACA) and anti β2 glycoprotein 1 (β2GP1) antibodies were tested on the BioFlash® platform (Inova Diagnostics, San Diego, CA, USA) with chemiluminescence methods; to assess positive result the manufacturer cut off 20 CU (Chemiluminescence Units) was used. IgG and IgM anti phosphatidylserine/prothrombin (PS/PT) antibodies were measured with a commercial ELISA kit on QUANTA-Lyser® 3000 (Inova Diagnostics, San Diego, CA, USA), using the manufacturer cut off (30 units). Biochemistry assays were tested on Atellica® Solution (Siemens Healthcare GmbH, Germany) and coagulation profile was assessed on CS-5100 System (Sysmex, Japan), using the manufacturer cut-offs.

### Statistical Analysis

The distribution of values of each parameter was assessed with Shapiro-Wilk test. The preliminary analysis of data revealed a non-normal distribution of values for most of them, so results were reported as median with 95% Confident Interval (95CI).

The statistical analysis was performed using Analyse-it® software version3.90.1 (Analyse-it® Software Ltd.; Leeds, UK). The study was conducted in accordance with the Helsinki Declaration and under the terms of all relevant local legislations. The investigation was based on pre-existing samples; approval was obtained from the local ethical committee.

### Funding source

None

## Results

The demographic characteristics of enrolled patients are summarized in *Table 1*.

**Table 1:** General characteristics of selected patients

|  |  |
| --- | --- |
| **Male/Female** (ratio) | 26/9 (2.9:1) |
| **Age (y) media** (range) | 73 (52-82) |
| **Days of hospitalization –median** (range) | 7 (2-28) |
| **Comorbidity (n°/total patients)** |  |
| *Informations not available* | 6/35 (17.1%) |
| *Hypertension* | 17/29 (58.6%) |
| *Cardiovascular Disease (different than hypertension)* | 10/29 (34.5%) |
| *Diabetes* | 8/29 (27.6%) |
| *Obesity* | 4/29 (13.8%) |
| *Kidney disease* | 5/29 (17.3%) |
| *Liver disease* | 3/29 (10.3%) |
| *Autoimmune disease* | 2/29 (6.9%) |
| *Hematological disease* | 2/29 (6.9%) |
| *Pulmonary disease* | 1/29 (3.4%) |
| *Gastrointestinal disease* | 1/29 (3.4%) |
| *No comorbidity* | 6/29 (20.7%) |
| **Autopsy Evidence**<br>(n° of patients with evidence of thrombosis of the small vessels ≥ 3 organs/total) | 35/35 (100%) |

The clinical and historical data were available for twenty-nine patients. 24/29 (82.8%) had one or more comorbidities. 3 patients (10.3%) were previously diagnosed for autoimmune diseases (2 Rheumatoid Arthritis and 1 Hashimoto Thyroiditis); only 1 patient was previously treated with oral anticoagulant and 7 patients with antiplatelet therapy. All the patients, but one, were treated for COVID-19 infection with a standard therapy composed by antibiotics, hydroxychloroquine and anti-retroviral drugs; in addition, 6 patients also received Tocilizumab, 1 Siltuximab and 6 Prednisone; 13 patients undergone a treatment with low molecular weight heparin (LMWH). A patient died before starting of therapy.

Laboratory features are showed in *Table 2*

**Table 2.** Main laboratory findings of patients *(CRP: C-Reactive Protein; LDH: Lactate Dehydrogenase PT: Prothrombin Time; INR: International Normalized Ratio; aPTT: activated Prothrombin Time; IL6: interleukin 6; PLT: platelet; AST: Aspartate transaminase; ALT: Alanine transaminase; hsTnI:* High-sensitivity cardiac troponin I; PCT: Procalcitonin)

| Test | N° samples (%) | range | Median<br>(95%CI) |
| --- | --- | --- | --- |
| CRP >1 mg/dL | 34/35(97.1) | 1.8-41.4 | 15.1<br>(11.9 to 21.2) |
| LDH >300 U/L | 32/35(91.4) | 338-17109 | 527<br>(448 to 687) |
| IL6 >3.4 pg/mL | 35/35(100) | 4.7-10000 | 129<br>(73 to 169) |
| D-Dimer >1000 ng/mL | 30/32(93.7) | 1367-35000 | 8508<br>(3531 to 23287) |
| PT (INR) >1.5 | 6/35(17.1) | 1.53-2.86 | 1.90<br>(1.53 to 2.86) |
| aPTT (Ratio) >1.25 | 17/33(51) | 1.29-2.32 | 1.49<br>(1.35 to 1.84) |
| PLT count <100 x10<sup>9</sup>/L | 2/35(5.7) | 36-67 | 52*<br>(nc) |
| Fibrinogen >450 mg/dL | 9/17(52.9) | 493-1193 | 748* |
|  |  |  | (584 to 912) |
| s-Creatinine >1.30<br>mg/dL | 20/35(57.1) | 1.35-7.22 | 2.76<br>(2.10 to 3.45) |
| AST >40 U/L | 25/34(73.5) | 41-7081 | 78<br>(65 to 102) |
| ALT >40 U/L | 20/35(57.1) | 41-2265 | 73<br>(57 to 232) |
| hsTnl > 53 ng/L | 9/13(69.2) | 174-11828 | 305<br>(174-11828) |
| PCT >10 ng/mL | 3/19(15.8) | 11.8-55.10 | 23.60*<br>(nc) |
| Ferritin >250 ng/mL | 29/34(85.3) | 296-16500 | 1489<br>(1116 to 2420) |

\*Mean value

Lactate dehydrogenase (LDH), C reactive protein (CRP) and interleukin 6 (IL-6) were markedly increased in all patients, except for one with slightly increased value for IL-6 alone, indicating an hyperinflammatory state.

The coagulation tests showed that all but one patient had increased value for D-Dimer, and in 12/32 patients (38.7%) the value was over 10000 ng/ml; prolonged prothrombin time (PT) and activate thromboplastin time (aPTT) were generally normal or slightly prolonged. Fibrinogen test was performed only in 17/35 patients, with increased values in 9/17(52.9%) but reduced in one. Only 6/17 (35.3%) patients fulfilled the DIC diagnostic criteria.

32/35 (91.4%) patients had evidence of kidney and/or hearth and/or liver damage.

10/35 patients (28.6%) were diagnosed for thromboembolic event with imaging techniques during the hospital stay: 4 patients with pulmonary embolism and 2 with cerebral ischemia.

The autopsy and the following microscopic evaluation on tissues samples demonstrated vascular involvement characterized by the presence of multiple recent microvascular and macrovascular thrombosis and the absence of microangiopathy with neutrophilic endothelial infiltrate in three or more organs in 35/35 (100%) patients, mainly lung, heart, liver and kidney.

Brain was evaluated in no autopsy due to bio-hazard security concerns.

*Table 3* summarizes the results of the aPL testing

**Table 3.** Results of anti phospholipid antibodies (aPL) testing

| **aPL findings (n=35)** | **Range**<br>**(CU, Units)** | **Median**<br>**(95%CI)** | **Positive**<br>**samples (%)** |
| --- | --- | --- | --- |
| Anti-cardiolipin IgG | 0.8-46.7 | 3.7<br>(2.9 to 4.4) | 1/35 (2.8) |
| Anti-cardiolipin IgM | 0.7-38.4 | 2.9<br>(1.9 to 4.0) | 2/35 (5.7) |
| Anti-cardiolipin IgA | 0.8-17.8 | 4.0<br>(2.5 to 5.2) | 0/35 (0) |
| Anti-β2 glycoprotein I IgG | 1.6-13.9 | 6.25*<br>(5.24 to 7.26) | 0/35 (0) |
| Anti-β2 glycoprotein I IgM | 0.1-9.7 | 1.2<br>(0.8 to 1.6) | 0/35 (0) |
| Anti-β2 glycoprotein I IgA | 0.0-10.3 | 0.3<br>(0.0 to 1.6) | 0/35 (0) |
| Phosphatidylserine/Prothrombin IgG | 5.5-34.4 | 8.5<br>(7.3 to 12.1) | 1/35 (2.8) |
| Phosphatidylserine/Prothrombin IgM | 1.2-50.4 | 7.6<br>(4.10 to 11.90) | 0/35 (0) |

\*Mean value

We found 3/35 (8.6%) positive patients: one for ACA IgG and two for ACA IgM but all the values were low (< 3X the cut off). No patients tested positive for ACA IgA neither for β2GP1 isotypes. 3/35 (8.6%) patients were positive for PS/PT, one for IgG and two for IgM, but values were less than 2X the cut off. No patients showed simultaneous positivity for ACA and PS/ PT. Between the three patients previously diagnosed with autoimmune diseases, only one tested positive for PS/PT IgG.

## Discussion

Venous and arterial thromboembolic phenomena are well described in a high percentage of COVID-19 patients, showing high levels of D-Dimer and, in some cases, prolonged coagulation tests, especially PT-INR [3–6, 12]; in those patients, the antithrombotic prophylaxis with LMWH or unfractionated heparin is recommended since it is associated with reduced mortality [13].

The thromboembolic disease in COVID 19 could have a multifactorial pathogenesis, as previously described [12, 14]. Hypoxia and prolonged immobilization are important factors, but the diffuse inflammatory state and the resulting “cytokine storm” seems to be the main triggers. The autopsies that have been conducted at the Pathology Department of the Papa Giovanni XXIII Hospital in Bergamo during the recent SARS-CoV-2 outbreak, highlighted a thrombotic damage in several organs that was clinically underestimated during the hospital stay. Several diseases such as DIC, sepsis, macrophage activated syndrome (MAS) and CAPS could be the underlying causes of such damages [15].

Recently some authors have been evidenced that the most COVID-19 patients meet the diagnostic criteria for DIC [13]; in our cohort, the appropriate assays and information were available for seventeen patients and only six (17.1%) meet the DIC criteria. From a clinical perspective, our patients did not show the typical features of DIC: platelet count and PT-INR were almost normal and fibrinogen values, when tested, were increased, possibly reflecting the hyperinflammation state; none of our patients except one showed hemorrhages and the histological findings consisting with both microvascular and macrovascular thrombosis did not fit with classical morphological feature of DIC.

Likewise, sepsis could be one of the likely causes of death in COVID-19 infection, but none of our patients fulfilled clinical criteria for this diagnosis and the histological post-mortem findings were not consistent with this assumption

MAS is another clinical condition that can be hypothetically related to the hypercoagulability state of COVID-19 patients: Carsana et al. [16] reported the presence of mainly monocyte/macrophage infiltration in lung biopsies highlighted with immunohistochemistry techniques, but normal platelet count, high level of fibrinogen and absence of hepato- and splenomegaly in our cohort of patients, exclude this hypothesis.

To the best of our knowledge, the presence of antiphospholipid antibodies in literature are reported only in 3 Chinese patients with multiple brain infarcts revealed with imaging techniques [17]. Our patients fulfilled the main clinical diagnostic criteria for CAPS described by Asherson RA et al. [9]: evidence of involvement in three or more organs; development of manifestations simultaneously or in less than a week, confirmation by histopathology of small vessel occlusion in at least one organ. Moreover, elevated levels of ferritin, like we found in our patients, were correlated with the most severe form of antiphospholipid syndrome (APS) such as CAPS [11]. Lastly, males are predominant in our COVID-19 cohort, instead aPL and correlated syndrome mainly affect female patients, but Rodriguez-Pinto et al. [10] reviewed the largest international CAPS cohort highlighting that in elderly patients the female male ratio is almost 1:1.

Based on these findings, to meet all the diagnostic criteria of CAPS, it was necessary to demonstrate the presence of aPL [10]. Unfortunately, in our cohort almost all the patients were negative for the aPL profile tested. Only 6/35 (17.1%) patients showed very low and not relevant antibodies levels but it is reported that slightly and transient increase of aPLs could be a common finding during any kind of infectious, whereas CAPS is always characterized by very high levels of autoantibodies [15]. Three conventional aPL markers are described in literature and used to diagnose CAPS: ACA, β2GPI and Lupus Anticoagulant (LAC). The main limit of our work it is that our patients were not tested for LAC due to the lack of post-mortem plasma samples. It is reported in literature a very high concordance between LAC and PS/PT testing [18, 19] and Litvinova et al. [20] reported that all CAPS patients are positive for both IgG and IgM PS/PT but only three of our patients were slightly positive for these tests. In particular, one out of two patients, found positive for PS/PT IgM was previously diagnosed with Hashimoto thyroiditis and was also positive for myeloperoxidase antibodies, thus suggesting a general activation of the immune system in an autoimmune-predisposed patient.

Another limit of our study is represented by the fact that some patients that were not tested for the complete biochemistry panel assays, mainly fibrinogen, which is a parameter essential for DIC scoring criteria or to contextualize other causes of hypercoagulation; this was due to incomplete knowledge of thromboembolic events and the hypercoagulability state of patients in the very first weeks of the pandemic.

In conclusion, the presence of systemic thromboembolic events in COVID-19 patients became quickly a well-known and established conditions, sometimes life-threatening. However, it is difficult to categorize those vascular events into a conventional disease: in our experience we did not find any significant association with anti-phospholipid antibodies. It is most likely that several factors contribute to trigger the hypercoagulability status and the thromboembolism but, on the basis our results, CAPS is probably not involved into the pathogenesis of these phenomena.

## Data Availability

NO

